# Wastewater surveillance for avian influenza: national patterns of detection and relationship with reported outbreaks and infections

**DOI:** 10.1101/2025.05.06.25327100

**Authors:** Marlene K Wolfe, Amanda L. Bidwell, Stephen P. Hilton, Alexandria B. Boehm

## Abstract

**Background:** Influenza A virus (IAV) is a major cause of morbidity and mortality globally, causing seasonal influenza in humans and infecting birds and some mammals. In 2024, IAV H5N1 highly pathogenic avian influenza (HPAI) in the United States moved into cattle. While the outbreak is currently of low risk to the public, there is an urgent need to monitor the disease and prevent spread.

**Methods:** We conducted a nationwide study evaluating the relationship between H5 hemagglutinin gene RNA concentrations in wastewater and reported outbreaks of IAV H5N1 in animals and humans. We utilized an H5-specific droplet digital RT-PCR test to quantify H5 RNA in wastewater in 40 states across the United States, and 1) examined the temporal association between outbreaks and wastewater detections and 2) utilized linear mixed models (LMM) to determine the relationship between measurements in wastewater and outbreak-related factors in the local area.

**Results:** We find that there is a significant temporal association between wastewater H5 detections and the incidence of outbreaks in poultry and wild birds, but not in cattle or with human infections. However, outbreaks tended to occur at the same time across populations - wild bird detections were also associated with H5N1 in herds, poultry, and humans. Utilizing a LMM, we find that for individual sites, there is a relationship between H5 measurements in wastewater and both poultry outbreaks and the presence of dairy industry locally, but that there was either no relationship or a negative relationship with H5 measurements and either combined systems that accept storm water or those with detection of H5 in wild birds.

**Conclusions:** The study highlights how wastewater monitoring can supplement traditional surveillance, providing vital data that reflects public health threats. The findings underscore the potential of scaled wastewater surveillance as a proactive tool in monitoring and managing future outbreaks.

## Introduction

Influenza A virus (IAV) is a leading cause of mortality and morbidity worldwide; the World Health Organization estimates that IAV causes up to 650,000 deaths and 1 billion cases each year.^1^ In the United States (US), it is responsible for an estimated annual 41 million infections, and 51,000 deaths, infecting approximately 8% of the US population.^2,3^ There is significant cost to associated illness as well; it is estimated to be responsible for $11.2 billion dollars in non-market and market economic losses each year in the US.^4,5^ This majority of this disease burden is due to the commonly circulating IAV human subtypes, however IAV subtypes are found in many animal reservoirs and zoonotic transmission from avian, swine, and other sources to humans as well as among other animals species is of concern^6^. The original source of both human and animal outbreaks of IAV in the past have been shown to originate in aquatic birds around the world,^6^ and the potential that IAV has to infect so many different species along with the ability of the segmented genome to readily undergo genetic reassortment means it has high pandemic potential^7^.

Influenza A H5N1 highly pathogenic avian influenza (HPAI or “avian influenza”) clade 2.3.4.4 is a subtype of IAV that is currently of global concern. It has affected over 100 million poultry since 2022, and in spring 2024, it made the interspecies jump to cattle^8,9^. As of April 2025, infections have been identified in 1,032 herds of cattle in 17 states in the US^10^, but risk of infection to humans remains low with 70 cases reported in humans^11^ primarily occurring among cattle and poultry workers, causing mild illness^12,13^. While the risk to the generally public is currently low, given that IAV can readily undergo genetic reassortment^7^ there is concern that H5N1 may acquire mutations that render it more transmissible to humans, particularly if co-infection with other more transmissible common human IAV subtypes occurs.

In spring of 2024, our team detected the H5 sequence of the hemagglutinin (HA) gene from the currently circulating avian influenza strain in wastewater samples from several wastewater treatment plants across the country where cattle were present in the vicinity^14^. As non-human animal waste does not typically enter the sanitary sewer system, direct inputs of animal waste to wastewater is an unlikely explanation for the H5 detection. Further investigation suggested that the H5 RNA marker in wastewater was likely from dairy waste discharged legally into the sanitary sewer from milk processing facilities, or through discarding of dairy products down household drains^14^. Follow-on modeling suggested that inputs of the H5 marker from wild birds or humans would be unrealistically high to account for concentrations of H5 marker typically observed in wastewater^15^. In a separate study, wastewater monitoring in Oregon (a state without cattle outbreaks) found that detections of H5 were most frequent in communities with wild bird habitats, but there was no significant association between detections and any animal outbreaks or dairy sources^16^. Given the apparent connection between wastewater detection of the H5 marker and presence of H5 in local dairies in our previous work, we expected data on H5 marker presence and concentration in wastewater would be useful for informing the response to the influenza A H5N1 outbreak - but as in the example in Oregon, there remains uncertainty about the multiple sources that may contribute to this signal.

With encouragement from public health collaborators from local and national agencies, we began prospectively measuring a marker of the H5 hemagglutinin gene in wastewater samples from wastewater treatment plants across the country in spring of 2024. Since then, we have measured the H5 marker continuously in wastewater, approximately three times per week, at 147 wastewater treatment plants in 40 states. In the current study, we present those data, investigate how concentrations of the H5 marker vary temporally and spatially, and how they relate with publicly available data on the number of infected cattle herds, poultry flocks, individual wildbirds and human cases in the US.

## Methods

### Wastewater sample collection for prospective study

The process for wastewater collection has been provided elsewhere^17^ previously and is summarized here. Samples were collected from 147 wastewater treatment plants (WWTPs) across 40 states between 15 May 2024 and 28 February 2025 approximately three times per week. A full description of the WWTP locations, as well as the number of samples and the time during which they were collected is provided in Appendix S1. WWTP staff provided either “grab” samples from the primary clarifier or 24-hour composite samples from the headworks (“influent”) (Appendix S1 provides sample type for each WWTP). Samples were then stored at 4°C and shipped to the laboratory on ice and processed to completion within 48 hours of receipt at the laboratory. We do not expect the time between sample collection and analysis to affect target quantification as our work and others^18–20^ show limited decay of the short length nucleic-acid targets over weeks at 4°C. A total of 18,023 samples were collected and analyzed for this study. Between 59 and 286 samples were tested per WWTP (median = 120 per WWTP).

### Pre-analytical processing and nucleic-acid extraction of wastewater samples

The pre-analytical processing and nucleic-acid extraction of the samples has been described previously in detail^21^ and is summarized here briefly. Wastewater solids are isolated from wastewater samples and re-suspended in a buffer at a low enough concentration so as to minimize inhibition, and homogenized. Samples were then subjected to nucleic-acid extraction and purification (Chemagic Viral DNA/RNA 300 Kit H96, PerkinElmer, Shelton, CT), and inhibitor removal (Zymo OneStep PCR Inhibitor Removal Kit, Irvine, CA). Dry weight was determined using another aliquot of solids and drying in an oven. Nucleic-acids from 6 to 10 aliquots of each sample were obtained and each used neat as template in 6 to 10 replicate droplet digital 1-step RT-PCR (dd-RT-PCR) reaction wells to measure the H5 marker. The process includes negative and positive extraction and PCR controls, as well as a measure of recovery as determined from spiking exogenous bovine coronavirus (BCoV). Further details are described in a protocol on protocols.io^22,23^ and in Boehm et al.^21^

### Droplet digital PCR

Concentrations of the H5 RNA marker in controls and samples were measured in multiplex droplet digital 1-step RT-PCR (dd-RT-PCR) reactions using an AutoDG Automated Droplet Generator (Bio-Rad, Hercules, CA), Mastercycler Pro (Eppendforf, Enfield, CT) thermocycler, and a QX600 Droplet Reader (Bio-Rad). The H5 assay was previous tested for sensitivity and specificity and shown to be highly specific to the currently circulating strain of H5N1^14^. The H5 assay was run using a probe-mixing multiplexing approach using probes labeled with ATTO590 in multiplex with other probe-based assays. Between 20 May 2024 and 16 July 2024, H5 was run as described in Boehm et al.^21^ in multiplex with assays for human metapneumovirus, *Candida auris*, enterovirus D68, and mpox clade II. Starting on 17 July 2024 and through the end of this study, the H5 assay was multiplexed with those same assays as well as assays for H1 and H3, variants of the influenza A HA gene. The change in assay mix was needed to respond to changing public health needs for influenza A monitoring in the United States, and did not affect performance of the H5 assay.

Digital droplet RT-PCR was performed on 20□μl samples from a 22□μl reaction volume, prepared using 5.5□μl template, mixed with 5.5□μl of One-Step RT-ddPCR Advanced kit for Probes (catalog no. 1863021; Bio-Rad), 2.2□μl reverse transcriptase (RT), 1.1□μl dithiothreitol (DTT), and primers and probes (Table S1) at a final concentration of 900□nM and 250□nM, respectively. Droplets were generated using the AutoDG Automated Droplet Generator (Bio-Rad).

PCR was performed using Mastercycler Pro with the following protocol: reverse transcription at 50□°C for 60□min, enzyme activation at 95°C for 5□min, 40 cycles with 1 cycle consisting of denaturation at 9□°C for 30□s and annealing and extension at 59□°C for 30□s, enzyme deactivation at 98□°C for 10□min, and then an indefinite hold at 4□°C. The ramp rate for temperature changes was set at 2□°C/s, and the final hold at 4□°C was performed for a minimum of 30□min to allow the droplets to stabilize.

Droplets were analyzed using the QX600 Droplet Reader (Bio-Rad). A well had to have over 10,000 droplets for inclusion in the analysis. Extraction and PCR positive and negative controls were run on each 96-well plate. Results from replicate wells were merged for analysis. Concentrations of the targets in wastewater samples are presented as copies per gram dry weight. For a sample to be scored as a positive, there had to be at least 3 positive droplets. The lowest measurable concentration is approximately 1000 copies/g dry weight (corresponds to three positive droplets). Errors are reported as standard deviations of the measurements as obtained from QX Manager Software (Bio-Rad, version 2.0).

### Number of livestock, poultry, and wildbird H5N1 infections

Livestock, poultry, and wildbird highly pathogenic avian influenza (HPAI) outbreak data was sourced from the US Department of Agriculture (USDA) Animal and Plant Health Inspection Service (APHIS) website on 6 March 2025^24^; the data are published and updated each weekday. The HPAI in cattle livestock data^10^ include the date of confirmed detection, the state where the detection occurred, and production type (backyard producers, dairy milking cows). HPAI poultry data^25^ include date of confirmed detection, state, county, and production type (commercial, backyard flock). HPAI wild bird data^26^ include information on state, county, sample collection date, detection date, HPAI strain, bird species, and sampling method.

### Dairy processing facilities within sewersheds

Surveys, email responses, and National Pollutant Discharge Elimination System (NPDES) permits were used to identify treatment plants that served dairy processing facilities. Whenever NPDES permits included addresses of dairy processing facilities, they were geocoded to obtain geographic coordinates using the tidygeocoder R package, and matched to polygons representing sewersheds using the sf package. The shapefiles for each sewershed were either provided by the WWTP, or were approximated using the zip codes that the utility serves. If dairy processing facilities identified with an NPDES permit were within one of these polygons, an online search was done to confirm that at least one of these facilities was operating during the study period and that the geocoded location was accurate. These sources only reflect some of the dairy manufacturing facilities in the US, and may exclude smaller and newer facilities in particular. However, they were selected as a reliable means to reflect which areas have higher levels of dairy production.

### Number of human H5 infections

Confirmed human influenza A H5N1 infections were sourced from the US Center for Disease Control and Prevention^11^ and downloaded on 7 March 2025.

### Statistics

Counts of newly infected herds/flocks, H5N1-positive wild birds, and H5N1 human infections were summarized weekly and aggregated across national, regional, and state spatial scales. Regional data were divided based on states included in the West, Northeast, South, and Midwest US regions as defined by the United States Census Bureau. Data on these detections were included for all states at the national and regional levels, including states for which there was no wastewater data available. For wastewater data, the proportion of wastewater samples tested that were positive for the H5 marker were calculated by Morbidity and Mortality Weekly Report (MMWR) week at each geographic scale. The peak in proportion of wastewater samples testing positive was calculated by MMWR week at the national and regional level and for all states with at least one wastewater site. All analysis was conducted in Rstudio using R version 4.1.2.

We used Kendall’s Tau to investigate the association between the number of detections of the H5 marker in wastewater and the detection of infected cattle herds, infected poultry flocks, infected wild birds, and infected humans on a weekly basis at the national, regional, and state levels. Kendall’s tau was used because detection counts and positivity were not normally distributed. A correlation matrix was generated for data aggregated across the nation, and for each state that had more than 10 detections of the H5 marker in wastewater (CA, CO, MI, NJ, TX). Kendall’s tau was also computed for the national data using the number of detections from wastewater, cattle herds, poultry flocks, wild birds and infected humans using only data from states with at least one site included in the study.

We used a linear mixed-effects model (LMM) to assess the relationship between the log_10_-transformed H5 concentration in wastewater and factors local to each utility that may plausibly be related to the presence of H5 in wastewater. Analysis was performed using the lmer function from the lme4 package in R version 4.1.2. Fixed effects tested for inclusion in the model included binary variables indicating the presence of dairy in the sewershed, the detection of any H5N1-infected in wild birds in the county, and whether a wastewater system was combined or separated; the number of reported poultry outbreaks in the county where the wastewater treatment plant is located was also included as a continuous variable, as well as the collection date of the sample. Variables were also tested in a linear model and the variance inflation factor (VIF) examined to confirm that there was no significant collinearity. Results below the limit of detection were replaced with approximately half the limit of detection, 500 cp/g, before log transformation. To account for repeated measurements at wastewater utilities, the model included a random intercept for the unique utility ID representing each individual wastewater site. This analysis was performed both for all sites at the national level, and for only sites in the state of California. California was chosen for this analysis because, unlike other geographic subsets of the data, there was both a large amount of data available, and variability in other variables included in the model across the dataset. California had the highest number of wastewater detections, infected cattle herds, human cases, and poultry outbreaks and among the highest number of wild birds detection among all states in which wastewater testing was performed.

## Results

### QA/QC

All the negative controls and positive controls were negative and positive for all analyses in this study. Results for recovery of bovine coronavirus were reported elsewhere and indicate near uniform recovery^21^. Additional details related to the environmental microbiology minimal information (EMMI) reporting guidelines, including the checklist, number of partitions, volume of partitions, copies per partition, and example fluorescent plots from the QX600 are reported in Boehm et al.^17^ Wastewater data are publicly available through the Stanford Digital Repository (https://purl.stanford.edu/bk537yf9916).

### H5 marker detection

The H5 marker was detected in 950 of 18,023 (5.27%) samples analyzed in the study. The marker was detected at least once at 69 of 147 (46.9%) WWTPs in 21 of 40 states (52%) (Figure S1). Concentrations were mostly low (57%), that is, within five times the approximately lower limit of detection (approximately 1000 cp/g) (Figure 1). However, at times, very high concentrations were observed; 48 (5%) samples had concentrations over 50,000 cp/g, and the highest measurement recorded was 1,025,122 cp/g. Of the 6 unique WWTPs that had concentrations over 50,000 copies/g measured in any sample, 4 sites (Turlock, CA, Los Angeles, CA, Dallas, TX, and Boise, ID) had dairy processing facilities discharging waste to the sanitary sewer system as reported by either facilities or federal databases, while the remaining 2 sites (Harrison, AR and Newark, NJ) did not (Appendix S1). One of these sites (Harrison, AR) has a sewer system that was described as combined, one (Turlock, CA) had at least one H5N1 poultry outbreak detected in the county during the study period, and one (Los Angeles, CA) had at least one detection of H5N1 in a wild bird during the study period.

**Figure 1.**
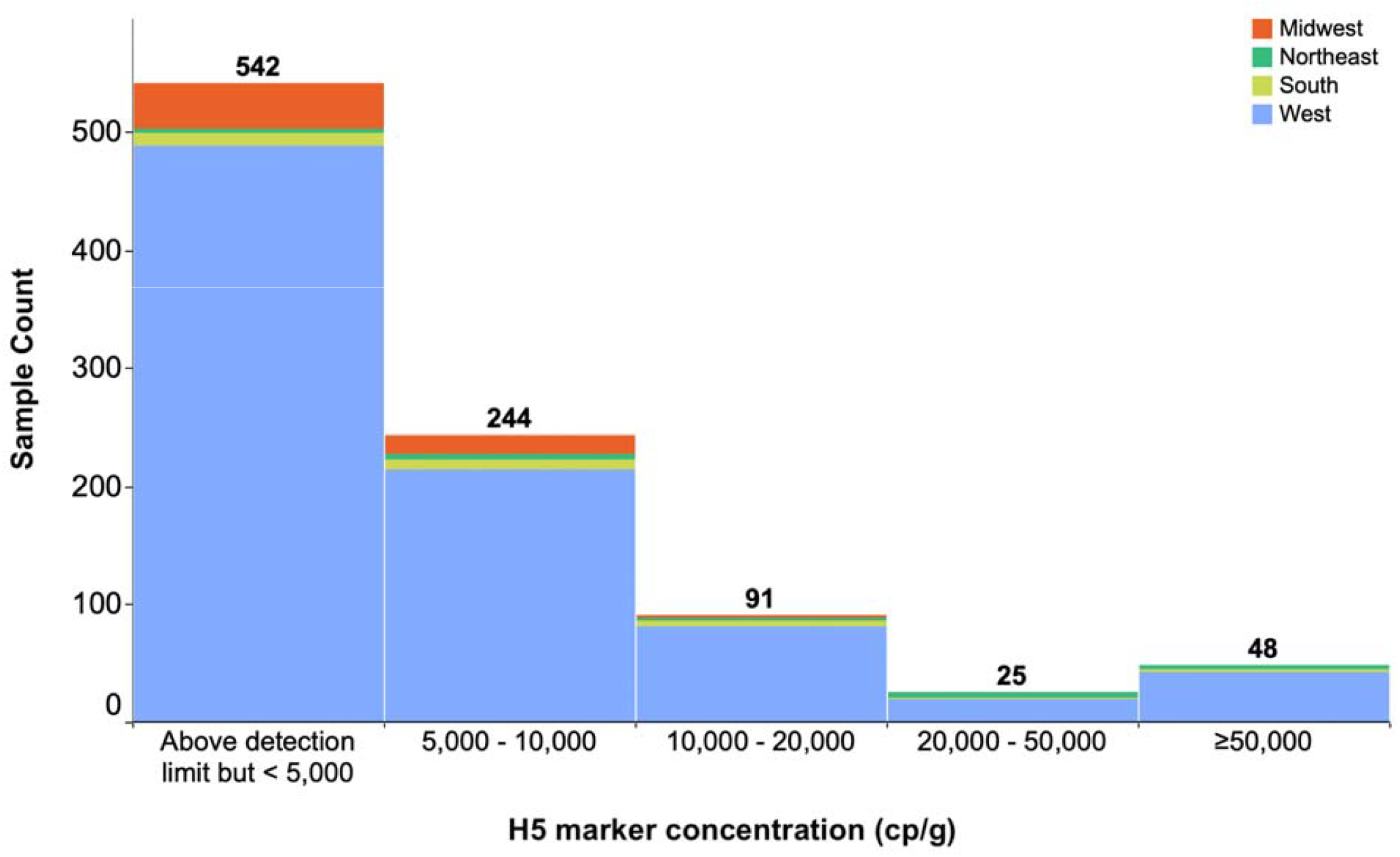
Histogram of measured concentrations of the H5 marker over the course of the study. The number above the column is the count of samples (out of 18,023) falling within the designated concentration range. Samples not included are those less than the lower detection limit (approximately 1000 copies per g dry weight (cp/g)).

### Timing and distribution of wastewater detections and outbreaks

For each state participating in the study, the date of the first detection of the H5 marker in wastewater from that state was compared to the first report of a cattle herd outbreak in that state (inclusive of first detections prior to the study period) (Figure 2). For 10 of the 21 states with a positive H5 wastewater test in this study, H5 marker was detected in wastewater and no cattle infections had been reported as of 1 March 2025. For 4 of the 21 states in this study, H5 marker was detected in wastewater before the first report of cattle infections (between 9 and 73 days early, median = 12 days). For 7 of the 21 states, the H5 marker was detected after the first report of cattle infections; 6 of these sites had already had a reported cattle infection prior to the start of the study (between 36 and 311 days early, median = 53 days).

**Figure 2.**
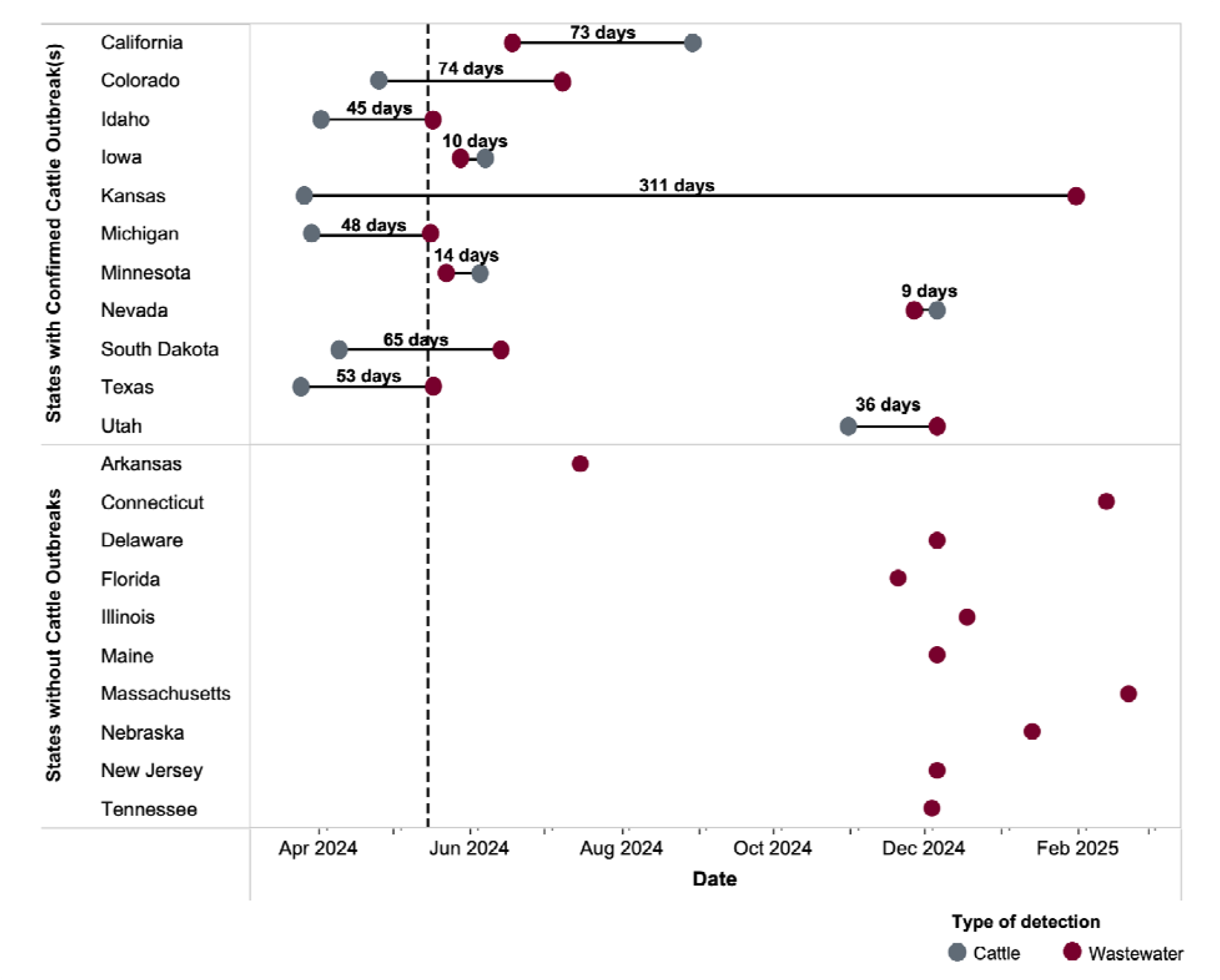
Time of the first wastewater detection of the H5 marker and cattle H5N1 detections (x-axis) by state (y-axis). The top panel shows states with confirmed cattle outbreaks and the bottom panel shows states without cattle outbreaks during the study duration. Dashed line represents the date monitoring for the H5 influenza marker began (15 May 2024). In the top panel, the duration above the lines indicates the time between detections.

The proportion of total samples positive for the H5 marker changed over time, reaching a peak nationally during the week beginning 22 December 2025 when 19% of samples nationally were positive for H5 (71 of 377 samples, Figure 3). Aggregated at the regional and state-level, the peak varied, occurring first in May 2024 in the South and Midwest, December 2024 in the West, and February 2025 in the Northeast (Table S2, Figure S2, S3). The number of H5N1 outbreaks and detections in aimals and people varied across the country; for example there were 869 herds that tested positive for H5N1 in the West during the study period, but only 45 in the midwest, 16 in the South, and 0 in the Northeast (Figure S4, Table S3).

**Figure 3.**
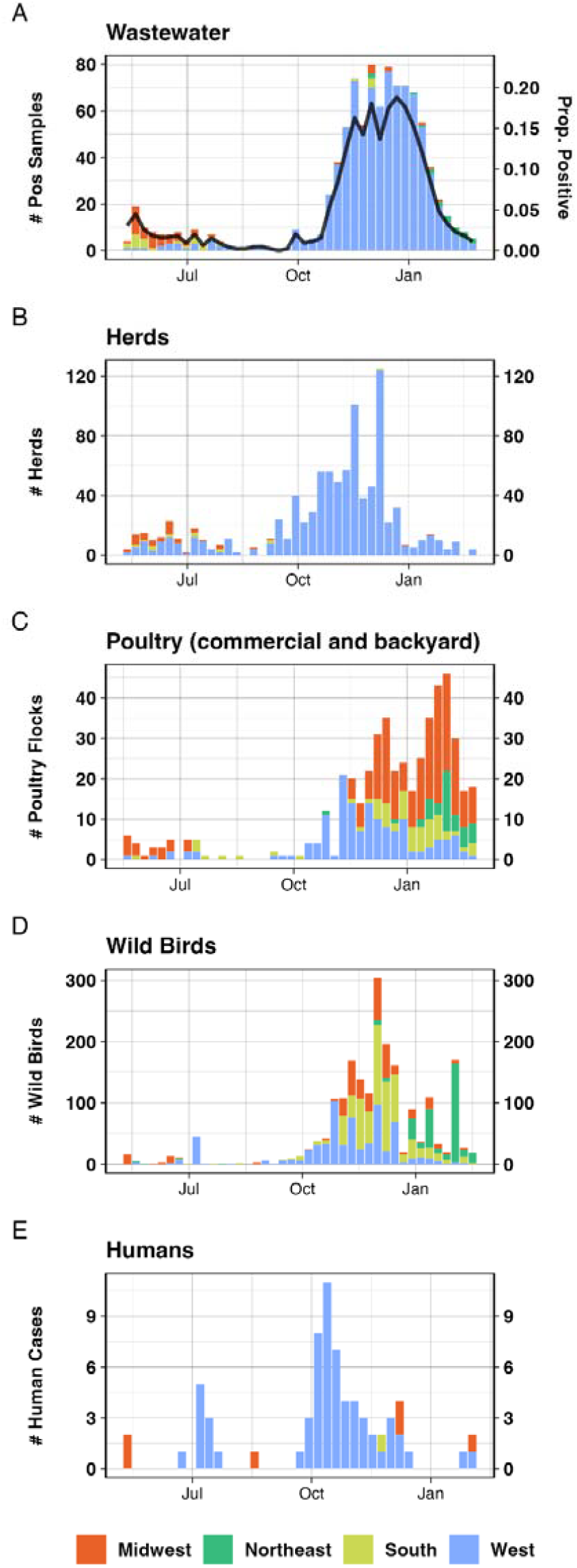
H5 influenza by MMWR week. (A) Weekly counts of influenza H5 marker detections in wastewater by week (left axis) and proportion of samples testing positive (right axis) starting 12 May 2024, MMWR 20 as a function of time compared to national H5 detections in (B) cattle herds, (C) poultry flocks, (D) individual wild birds, and (E) human cases of H5N1 infections. The location of the H5 detections are represented by region; the Midwest is in orange, Northeast is in green, South is in yellow, and West is in blue.

### Association between wastewater positivity and outbreaks over time

Aggregated weekly across the nation, the proportion of wastewater samples positive for the H5 marker correlated significantly and positively with the total number of new poultry outbreaks and wild bird outbreaks (Kendall’s tau = 0.53, p = 1.26*10^−5^ and tau = 0.55, p = 3.74*10^−6^ respectively), but not with the number of new outbreaks in cattle herds or number of human infections (tau = 0.29, p = 0.08 and tau = 0.13, p = 1.0 respectively, Figure 4, Table S3). Wild birds were also significantly associated with occurrence of H5N1 outbreaks in herds and poultry as well as human cases, nationally. This analysis was repeated using only data from the 40 states with at least one wastewater site in the study, and the results did not change substantially.

**Figure 4.**
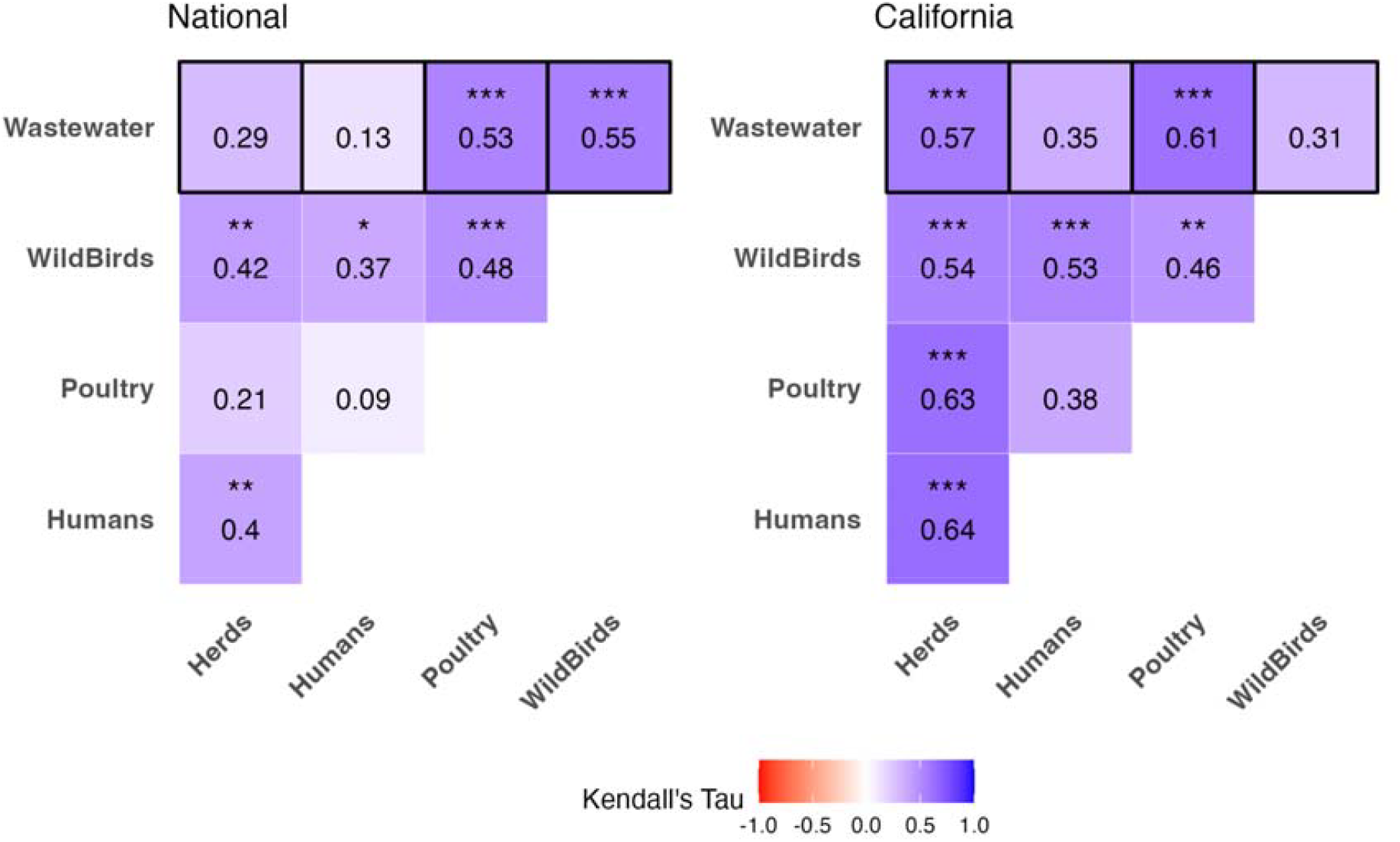
Correlation matrix of association between wastewater positivity and outbreaks over time at the National program level and for California. P-value <0.05 = *, <0.01 = **, < 0.001 = ***.

At the state level, the proportion of positive wastewater samples in California was significantly associated with the number of cattle herd outbreaks (tau = 0.57, p = 1.22*10^−5^) and poultry outbreaks (tau = 0.61, p = 6.0*10^−6^) per week; there were also significant associations between the occurrence of outbreaks in herds with outbreaks/ infections in wild birds, poultry, and humans, and wild bird outbreaks were also associated with outbreaks in poultry and detection in humans. Associations were also computed for the other 4 states with more than 10 detections of H5 in a WWTP during the study period (CO, MI, NJ, TX). Results varied by state, with significant associations between dairy herd outbreaks and wastewater detection of H5 in TX and CO; full analysis is included in the SI (Table S3, Figure S4).

### Relationship between local sources of H5N1 and H5 concentration in wastewater

We used a LMM to examine the relationship between the log_10_-transformed H5 marker concentration in wastewater and factors related to local sources of H5 at each utility. We found that, using the entire dataset of sites across the US, the presence of dairy facilities in the sewershed and the number of poultry outbreaks in the county associated with the sewershed were significantly associated with higher log_10_ concentrations of H5 observed in the local wastewater (estimate = 0.06, p = 1.08*10^−3^ and estimate = 0.02, p = 4.61*10^−8^, respectively; Table 1, Table S4) in a model fitting a total of 18,023 observations from 147 sites. Time (Collection Date) was included as a fixed effect, as inclusion improved the model as assessed by Bayesian Information Criterion (BIC) score. BIC score is a model selection tool that assesses the performance of a model and helps avoid overfitting; lower BIC scores indicate better performance. Site level random effects accounted for a moderate amount of the total variance in H5 concentration (12.6%).

**Table 1.** Results from a linear mixed model describing the relationship between wastewater concentrations of H5 RNA and characteristics of wastewater utilities, with site included as a random effect. P-value <0.05 = *, <0.01 = **, < 0.001 = ***. Full results are available in Table S4.

|  | National | West | South | Midwest | Northeast | California |
| --- | --- | --- | --- | --- | --- | --- |
| Dairy in sewershed | 0.06** | 0.14** | 0.02** | -0.01 | -0.01 | 0.15* |
| # of poultry outbreaks in county | 0.02*** | 0.02** | 0.00 | 0.00 | 0.01 | 0.02* |
| Wild bird detection in county | -0.01 | -0.05 | 0.00 | 0.00 | -0.02 | -0.14* |
| Separated System | 0.00 | -0.03 | 0.00 | 0.01 | 0.02 | 0.05 |
| Collection Date | 0.00*** | 0.00*** | 0.00*** | 0.00*** | 0.00*** | 0.00*** |

The same model was run at the regional level and for sites in the state of California, which had the highest number of wastewater detections, infected cattle herds, human cases, and poultry outbreaks and among the highest number of wild birds detection among all states in which wastewater testing was performed. In California, results indicated that the presence of dairy operations in the sewershed and the number of poultry outbreaks were significantly associated with higher H5 concentrations (estimate = 0.15, p = 0.019 and estimate = 0.02, p = 0.015, respectively; Table 1, Table S4), but that the presence of a wild bird detection in the county was associated with a significantly lower concentrations of H5 (estimate = −0.14, p = 0.031). Site level random effects accounted for 9.7% of the total variance in H5 concentration observed in California sites.

## Discussion

This study demonstrates how nationally scaled wastewater monitoring can supplement available data about H5N1 infections in specific animal and human populations, reflecting the outbreak as it has spread across the United States. We demonstrate that nationally, there is a significant association between the timing of H5N1 outbreaks as measured across different populations. In California, where the outbreaks were concentrated for much of the study period, the weekly number of outbreaks in cattle herds was significantly associated with weekly detections of H5N1 in wastewater as well as identification of infections in all studied populations: poultry, wild birds, and humans. This pattern demonstrates temporal coherence across outbreaks in different species that was also reflected in wastewater, but does not help establish the source of the H5 that was detected in wastewater.

To better understand likely sources of H5 in wastewater, we looked at the relationship between possible H5N1 sources and wastewater concentrations of H5 in 18,023 samples from 40 states over a period of approximately 9 months. Because of the nature of wastewater testing - namely, that an aggregated sample that represents many mixed, anonymous sources provides community-level information - it is difficult to determine the likely source of disease markers that appear in a sample. Our analysis showed a significant relationship between H5 concentrations in wastewater and both the presence of dairy facilities and the number of poultry outbreaks in the area surrounding the wastewater utility. This evidence supports the hypothesis that cattle and in some cases poultry are the most likely contributors of H5N1 that is detected in wastewater, previously established in our work measuring H5 in dairy discharge upstream of wastewater utilities^14^ and modeling the impact of different animal contributions to wastewater^15^.

Because of difficulty in demonstrating the source of H5 influenza subtypes in wastewater, there is concern that wild birds and combined sewer systems that accept environmental inputs are a source of H5N1 in wastewater, making this information less useful as a way of tracking outbreaks among domestic herds and flocks. Our results show that there was no significant relationship between either wild bird detections of H5N1 or sewer system type and H5 concentrations in wastewater when all sites nationally were included. When the analysis was limited to California sites, there was a negative relationship between wild bird detections of H5N1 and H5 concentrations in wastewater. While each situation is unique and depends on the context of local outbreaks and sewer system design and drainage, these results suggest that waste from wild birds is an unlikely driver of H5 concentrations in wastewater. This conclusion is supported by recent modeling work from our team^15^, which also suggests that the volume of bird feces entering the sewer system would need to be extraordinarily high to produce the results that have been observed in wastewater.

It has previously been suggested that identifying particularly high measured concentrations of influenza A RNA in wastewater, or detections of influenza A RNA in wastewater (using a general influenza A marker like one within the M gene) outside of the usual flu season in North America, might be useful indicators of local of H5N1 animal outbreaks^27^. However, during almost a year of wastewater monitoring, the majority of detections of H5N1 were at low to moderate levels, and the majority of the detections occurred during what is also the typical flu season from October - January^21^. These results suggest that it is important to monitor specifically for an H5 marker if H5 subtypes are of interest, rather than relying on a general marker for influenza A to identify outbreak areas.

There are limitations both to H5 monitoring in wastewater using this approach, and the comparative analysis presented in this study. We utilized surveillance data from a number of systems that each have their own limitations. Data on outbreaks in cattle and poultry and infections in wild birds and humans are limited by the systems that collect such data, and there have been substantial challenges in identifying and tracking infected cattle herds in particular and especially at the begin of the outbreaks in the US before systems such as bulk milk tank monitoring were implemented. Data on detections in wild birds and humans depends on the identification and testing of individuals who are infected, and for both populations it is unlikely that all infections have been identified. Finally, we have matched data on human and wild bird infections and poultry outbreaks to wastewater utilities based on county. This may or may not mean that there is a direct physical connection between these infections and the sewer systems. We have also based our analysis on the presence of any known dairy in the sewershed, as it is infeasible to identify dairy sources that are known to contain H5N1 RNA. However, this is a conservative approach to establishing a connection between contaminated dairy and H5 in wastewater, suggesting that the relationship identified in this analysis is robust. More information on the presence of H5 in milk within sewersheds would help characterize this relationship further, however this is outside the scope of current data collection and programs and it would be a challenge to acquire such data.

There are still many questions to answer about H5 in wastewater, and further characterization of the virus by genotyping or sequencing-based methods may help illuminate the patterns of these outbreaks and the sources of H5 shedding producing the observed signal. This work demonstrates that H5 monitoring in wastewater produces valuable data and can be scaled to large, country-wide systems in a way that provides data that is associated with outbreaks of importance to public health.

## Data Availability

All wastewater data are publicly available through the Stanford Digital Repository (https://purl.stanford.edu/bk537yf9916)

## Acknowledgments

We thank the participating wastewater treatment plants for their samples for the project. This work was supported by a gift to ABB from the Sergey Brin Family Foundation.

## Notes

### Competing Interest Statement

The authors have declared no competing interest.

### Author Declarations

The study used ONLY openly available human data that were originally located at https://www.cdc.gov/bird-flu/h5-monitoring/index.html

