## Supplementary Information for "Wastewater surveillance for avian influenza: national patterns of detection and relationship with reported outbreaks and infections"

**Tables	2**

Table S1. H5 Primers and probes, as previously reported in Wolfe et al1	2

Table S2. Peak wastewater positivity in terms of proportion of samples positive for H5 in a given week by location.	3

Table S3. Total counts of positive herds, humans, poultry, wastewater samples, and wild birds recorded over the duration of the study, by state	4

Table S3. Kendall’s tau values	6

Table S4. All LMM results	8

**Figures	10**

Figure S1. Map of the wastewater treatment plants (WWTPs) in this study.	11

Figure S2. Wastewater sample positivity by region over time.	12

Figure S3. Wastewater samples positivity by state.	13

Figure S4. Total counts of positive herds, humans, poultry, wastewater samples, and wild birds recorded over the duration of the study, by region	14

Figure S5. Correlation matrix for states with more than 10 detections of H5 in a WWTP during the study period (CA, CO, MI, NJ, TX).	15

### Tables

##### **Table S1.** H5 Primers and probes, as previously reported in Wolfe et al^1^

| Forward Primer | TATAGARGGAGGATGGCAGG |
| --- | --- |
| Reverse Primer | ACDGCCTCAAAYTGAGTGTT |
| Probe | AGGGGAGTGGKTACGCTGCRGAC |

##### **Table S2**. Peak wastewater positivity in terms of proportion of samples positive for H5 in a given week by location.

No positive samples during the study period were observed in Alabama, Alaska, Georgia, Hawaii, Indiana, Kentucky, Louisiana, Maryland, Mississippi, New Hampshire, New York, North Carolina, Ohio, Pennsylvania, Vermont, Virginia, Washington, West Virginia, or Wisconsin.

| **Region** | **Week start of wastewater peak** | **Proportion Positive** |
| --- | --- | --- |
| **National** | 12/22/2024 | 0.19 |
| **Regions** |  |  |
| South | 5/12/2024 | 0.05 |
| Midwest | 5/19/2024 | 0.12 |
| West | 12/22/2024 | 0.55 |
| Northeast | 02/16/2025 | 0.06 |
| **States** |  |  |
| Idaho | 5/12/2024 | 0.33 |
| Texas | 5/12/2024 | 0.22 |
| Michigan | 5/19/2024 | 0.50 |
| Minnesota | 5/19/2024 | 0.25 |
| South Dakota | 6/16/2024 | 0.67 |
| Arkansas | 7/14/2024 | 0.33 |
| Colorado | 7/21/2024 | 0.83 |
| Florida | 11/17/2024 | 0.03 |
| Nevada | 11/24/2024 | 0.33 |
| Delaware | 12/1/2024 | 0.33 |
| Iowa | 12/1/2024 | 0.27 |
| Utah | 12/1/2024 | 0.20 |
| Tennessee | 12/1/2024 | 0.17 |
| Maine | 12/1/2024 | 0.11 |
| California | 12/15/2024 | 0.70 |
| Illinois | 12/15/2024 | 0.17 |
| Nebraska | 01/12/2025 | 0.17 |
| Kansas | 01/26/2025 | 0.07 |
| New Jersey | 02/02/2025 | 0.17 |
| Connecticut | 02/09/2025 | 0.33 |
| Massachusetts | 02/16/2025 | 0.20 |

##### **Table S3.** Total counts of positive herds, humans, poultry, wastewater samples, and wild birds recorded over the duration of the study, by state

| **State/Region** | **Herds** | **Poultry** | **Wild Birds** | **Humans** | **Wastewater** |
| --- | --- | --- | --- | --- | --- |
| Midwest | 45 | 228 | 392 | 6 | 57 |
| Northeast | 0 | 41 | 331 | 0 | 19 |
| South | 16 | 64 | 676 | 1 | 28 |
| West | 869 | 148 | 670 | 62 | 846 |
| Alabama | 0 | 2 | 19 | 0 | 0 |
| Alaska | 0 | 2 | 62 | 0 | 0 |
| Arizona | 1 | 5 | 74 | 0 | 0 |
| Arkansas | 0 | 9 | 30 | 0 | 2 |
| California | 748 | 77 | 119 | 38 | 821 |
| Colorado | 62 | 11 | 54 | 10 | 12 |
| Connecticut | 0 | 3 | 19 | 0 | 1 |
| Delaware | 0 | 4 | 6 | 0 | 1 |
| Florida | 0 | 16 | 168 | 0 | 2 |
| Georgia | 0 | 3 | 17 | 0 | 0 |
| Hawaii | 0 | 2 | 9 | 0 | 0 |
| Idaho | 32 | 18 | 15 | 0 | 10 |
| Illinois | 0 | 4 | 48 | 0 | 1 |
| Indiana | 0 | 20 | 14 | 0 | 0 |
| Iowa | 13 | 15 | 96 | 1 | 10 |
| Kansas | 0 | 10 | 58 | 0 | 1 |
| Kentucky | 0 | 0 | 38 | 0 | 0 |
| Louisiana | 0 | 3 | 58 | 1 | 0 |
| Maine | 0 | 2 | 117 | 0 | 2 |
| Maryland | 0 | 6 | 9 | 0 | 0 |
| Massachusetts | 0 | 2 | 12 | 0 | 1 |
| Michigan | 17 | 12 | 42 | 2 | 30 |
| Minnesota | 9 | 30 | 15 | 0 | 7 |
| Mississippi | 0 | 2 | 111 | 0 | 0 |
| Missouri | 0 | 30 | 60 | 1 | 0 |
| Montana | 0 | 2 | 20 | 0 | 0 |
| Nebraska | 0 | 8 | 12 | 0 | 1 |
| Nevada | 10 | 1 | 16 | 1 | 1 |
| New Hampshire | 0 | 0 | 92 | 0 | 0 |
| New Jersey | 0 | 1 | 8 | 0 | 15 |
| New Mexico | 1 | 0 | 16 | 0 | 0 |
| New York | 0 | 13 | 20 | 0 | 0 |
| North Carolina | 0 | 4 | 18 | 0 | 0 |
| North Dakota | 0 | 4 | 11 | 0 | 0 |
| Ohio | 0 | 72 | 12 | 1 | 0 |
| Oklahoma | 2 | 5 | 47 | 0 | 0 |
| Oregon | 1 | 10 | 49 | 1 | 0 |
| Pennsylvania | 0 | 17 | 46 | 0 | 0 |
| Rhode Island | 0 | 1 | 6 | 0 | 0 |
| South Carolina | 0 | 2 | 30 | 0 | 0 |
| South Dakota | 6 | 20 | 7 | 0 | 7 |
| Tennessee | 0 | 3 | 29 | 0 | 1 |
| Texas | 14 | 2 | 61 | 0 | 22 |
| Utah | 13 | 7 | 93 | 0 | 2 |
| Vermont | 0 | 2 | 11 | 0 | 0 |
| Virginia | 0 | 2 | 29 | 0 | 0 |
| Washington | 0 | 9 | 130 | 11 | 0 |
| West Virginia | 0 | 1 | 6 | 0 | 0 |
| Wisconsin | 0 | 3 | 17 | 1 | 0 |
| Wyoming | 1 | 4 | 13 | 1 | 0 |

##### **Table S3.** Kendall’s tau values nationally and for states with more than 10 detections of H5 in a WWTP during the study period (CA, CO, MI, NJ, TX). Results are summarized in heatmaps in Figure 4 in the main text and Figure S5.

P-value <0.05 = *, <0.01 = **, < 0.001 = ***.

| **Area** | **Variable 1** | **Variable 2** | **Tau** | **S** | **p-value** | **Adjust p-value** | **Signif** |
| --- | --- | --- | --- | --- | --- | --- | --- |
| National | Herds | Poultry | 0.21 | 1.91 | 5.56E-02 | 5.56E-01 |  |
| National | Humans | Poultry | 0.09 | 0.72 | 4.74E-01 | 1.00E+00 |  |
| National | Herds | WildBirds | 0.42 | 3.82 | 1.35E-04 | 1.35E-03 | ** |
| National | Poultry | WildBirds | 0.48 | 4.30 | 1.74E-05 | 1.74E-04 | *** |
| National | Humans | WildBirds | 0.37 | 3.12 | 1.79E-03 | 1.79E-02 | * |
| National | Herds | Humans | 0.40 | 3.33 | 8.80E-04 | 8.80E-03 | ** |
| National | Herds | Wastewater | 0.29 | 2.67 | 7.55E-03 | 7.55E-02 |  |
| National | Poultry | Wastewater | 0.53 | 4.85 | 1.26E-06 | 1.26E-05 | *** |
| National | WildBirds | Wastewater | 0.55 | 5.08 | 3.74E-07 | 3.74E-06 | *** |
| National | Humans | Wastewater | 0.13 | 1.08 | 2.79E-01 | 1.00E+00 |  |
| California | Herds | Poultry | 0.63 | 5.00 | 5.71E-07 | 5.71E-06 | *** |
| California | Humans | Poultry | 0.38 | 2.77 | 5.53E-03 | 5.53E-02 |  |
| California | Herds | WildBirds | 0.54 | 4.40 | 1.06E-05 | 1.06E-04 | *** |
| California | Poultry | WildBirds | 0.46 | 3.57 | 3.51E-04 | 3.51E-03 | ** |
| California | Humans | WildBirds | 0.53 | 4.00 | 6.26E-05 | 6.26E-04 | *** |
| California | Herds | Humans | 0.64 | 4.94 | 7.73E-07 | 7.73E-06 | *** |
| California | Herds | Wastewater | 0.57 | 4.85 | 1.22E-06 | 1.22E-05 | *** |
| California | Poultry | Wastewater | 0.61 | 4.99 | 6.00E-07 | 6.00E-06 | *** |
| California | WildBirds | Wastewater | 0.31 | 2.55 | 1.06E-02 | 1.06E-01 |  |
| California | Humans | Wastewater | 0.35 | 2.79 | 5.19E-03 | 5.19E-02 |  |
| Colorado | Herds | Poultry | 0.17 | 1.15 | 2.49E-01 | 1.00E+00 |  |
| Colorado | Humans | Poultry | 0.47 | 3.07 | 2.16E-03 | 2.16E-02 | * |
| Colorado | Herds | WildBirds | -0.05 | -0.31 | 7.55E-01 | 1.00E+00 |  |
| Colorado | Poultry | WildBirds | 0.26 | 1.68 | 9.33E-02 | 9.33E-01 |  |
| Colorado | Humans | WildBirds | 0.13 | 0.87 | 3.84E-01 | 1.00E+00 |  |
| Colorado | Herds | Humans | 0.58 | 3.89 | 1.00E-04 | 1.00E-03 | ** |
| Colorado | Herds | Wastewater | 0.48 | 3.28 | 1.02E-03 | 1.02E-02 | * |
| Colorado | Poultry | Wastewater | -0.06 | -0.41 | 6.85E-01 | 1.00E+00 |  |
| Colorado | WildBirds | Wastewater | 0.02 | 0.15 | 8.79E-01 | 1.00E+00 |  |
| Colorado | Humans | Wastewater | 0.32 | 2.07 | 3.89E-02 | 3.89E-01 |  |
| Michigan | Herds | Poultry | -0.08 | -0.54 | 5.87E-01 | 1.00E+00 |  |
| Michigan | Humans | Poultry | -0.07 | -0.45 | 6.56E-01 | 1.00E+00 |  |
| Michigan | Herds | WildBirds | 0.23 | 1.52 | 1.29E-01 | 1.00E+00 |  |
| Michigan | Poultry | WildBirds | 0.16 | 1.11 | 2.69E-01 | 1.00E+00 |  |
| Michigan | Humans | WildBirds | 0.42 | 2.78 | 5.45E-03 | 5.45E-02 |  |
| Michigan | Herds | Humans | 0.26 | 1.67 | 9.55E-02 | 9.55E-01 |  |
| Michigan | Herds | Wastewater | 0.36 | 2.46 | 1.38E-02 | 1.38E-01 |  |
| Michigan | Poultry | Wastewater | -0.20 | -1.37 | 1.69E-01 | 1.00E+00 |  |
| Michigan | WildBirds | Wastewater | 0.00 | 0.00 | 1.00E+00 | 1.00E+00 |  |
| Michigan | Humans | Wastewater | 0.32 | 2.11 | 3.51E-02 | 3.51E-01 |  |
| New Jersey | Herds | Poultry | NA | NA | NA | NA |  |
| New Jersey | Humans | Poultry | NA | NA | NA | NA |  |
| New Jersey | Herds | WildBirds | NA | NA | NA | NA |  |
| New Jersey | Poultry | WildBirds | -0.04 | -0.28 | 7.82E-01 | 1.00E+00 |  |
| New Jersey | Humans | WildBirds | NA | NA | NA | NA |  |
| New Jersey | Herds | Humans | NA | NA | NA | NA |  |
| New Jersey | Herds | Wastewater | NA | NA | NA | NA |  |
| New Jersey | Poultry | Wastewater | 0.34 | 2.29 | 2.21E-02 | 6.64E-02 |  |
| New Jersey | WildBirds | Wastewater | -0.13 | -0.85 | 3.93E-01 | 1.00E+00 |  |
| New Jersey | Humans | Wastewater | NA | NA | NA | NA |  |
| Texas | Herds | Poultry | -0.07 | -0.48 | 6.30E-01 | 1.00E+00 |  |
| Texas | Humans | Poultry | NA | NA | NA | NA |  |
| Texas | Herds | WildBirds | -0.09 | -0.60 | 5.46E-01 | 1.00E+00 |  |
| Texas | Poultry | WildBirds | 0.25 | 1.69 | 9.18E-02 | 5.51E-01 |  |
| Texas | Humans | WildBirds | NA | NA | NA | NA |  |
| Texas | Herds | Humans | NA | NA | NA | NA |  |
| Texas | Herds | Wastewater | 0.40 | 2.76 | 5.80E-03 | 3.48E-02 | * |
| Texas | Poultry | Wastewater | -0.09 | -0.59 | 5.57E-01 | 1.00E+00 |  |
| Texas | WildBirds | Wastewater | -0.26 | -1.81 | 7.03E-02 | 4.22E-01 |  |
| Texas | Humans | Wastewater | NA | NA | NA | NA |  |

##### **Table S4.** Full results from a linear mixed model describing the relationship between wastewater concentrations of H5 RNA and characteristics of wastewater utilities, with site included as a random effect. These results are summarized in Table 1 in the full text.

| **Group** | **Variable** | **Estimate** | **Standard Error** | **df** | **t-value** | **p-value** |
| --- | --- | --- | --- | --- | --- | --- |
| **National** | (Intercept) | -4.63 | 0.41 | 17938.83 | -11.37 | 0.0000 |
| **National** | Dairy in sewershed | 0.06 | 0.02 | 140.02 | 3.34 | 0.0011 |
| **National** | # of poultry outbreaks in county | 0.02 | 0.00 | 151.61 | 5.76 | 0.0000 |
| **National** | Wild bird detection in county | -0.01 | 0.02 | 187.46 | -0.41 | 0.6846 |
| **National** | Separated System | 0.00 | 0.02 | 141.61 | -0.09 | 0.9323 |
| **National** | Collection Date | 0.00 | 0.00 | 17880.56 | 18.07 | 0.0000 |
| **South** | (Intercept) | 4.29 | 0.28 | 5955.03 | 15.37 | 0.0000 |
| **South** | Dairy in sewershed | 0.02 | 0.01 | 49.07 | 3.34 | 0.0016 |
| **South** | # of poultry outbreaks in county | 0.00 | 0.00 | 64.86 | -0.43 | 0.6667 |
| **South** | Wild bird detection in county | 0.00 | 0.01 | 46.66 | -0.01 | 0.9938 |
| **South** | Separated System | 0.00 | 0.01 | 50.16 | -0.19 | 0.8482 |
| **South** | Collection Date | 0.00 | 0.00 | 5947.97 | -5.70 | 0.0000 |
| **West** | (Intercept) | -22.01 | 1.12 | 5846.01 | -19.73 | 0.0000 |
| **West** | Dairy in sewershed | 0.14 | 0.05 | 34.84 | 2.77 | 0.0090 |
| **West** | # of poultry outbreaks in county | 0.02 | 0.01 | 35.25 | 3.46 | 0.0014 |
| **West** | Wild bird detection in county | -0.05 | 0.04 | 35.22 | -1.21 | 0.2362 |
| **West** | Separated System | -0.03 | 0.06 | 34.54 | -0.46 | 0.6513 |
| **West** | Collection Date | 0.00 | 0.00 | 5822.65 | 22.28 | 0.0000 |
| **Midwest** | (Intercept) | 5.92 | 0.42 | 3942.65 | 14.12 | 0.0000 |
| **Midwest** | Dairy in sewershed | -0.01 | 0.01 | 27.55 | -0.58 | 0.5653 |
| **Midwest** | # of poultry outbreaks in county | 0.00 | 0.00 | 27.41 | -0.28 | 0.7798 |
| **Midwest** | Wild bird detection in county | 0.00 | 0.01 | 27.66 | 0.20 | 0.8466 |
| **Midwest** | Separated System | 0.01 | 0.01 | 27.69 | 1.51 | 0.1427 |
| **Midwest** | Collection Date | 0.00 | 0.00 | 3940.55 | -7.68 | 0.0000 |
| **Northeast** | (Intercept) | -1.57 | 0.73 | 2184.14 | -2.14 | 0.0324 |
| **Northeast** | Dairy in sewershed | -0.01 | 0.03 | 14.65 | -0.26 | 0.7985 |
| **Northeast** | # of poultry outbreaks in county | 0.01 | 0.03 | 14.94 | 0.31 | 0.7641 |
| **Northeast** | Wild bird detection in county | -0.02 | 0.03 | 14.68 | -0.59 | 0.5650 |
| **Northeast** | Separated System | 0.02 | 0.03 | 14.71 | 0.69 | 0.5004 |
| **Northeast** | Collection Date | 0.00 | 0.00 | 2179.74 | 5.83 | 0.0000 |
| **California** | (Intercept) | -31.29 | 1.40 | 4459.32 | -22.42 | 0.0000 |
| **California** | Dairy in sewershed | 0.15 | 0.06 | 22.74 | 2.53 | 0.0188 |
| **California** | # of poultry outbreaks in county | 0.02 | 0.01 | 23.17 | 2.63 | 0.0149 |
| **California** | Wild bird detection in county | -0.14 | 0.06 | 23.01 | -2.29 | 0.0315 |
| **California** | Separated System | 0.05 | 0.07 | 22.65 | 0.72 | 0.4802 |
| **California** | Collection Date | 0.00 | 0.00 | 4445.35 | 24.48 | 0.0000 |

### Figures

##### Figure S1. Map of the wastewater treatment plants (WWTPs) in this study.

States where the H5 influenza marker was detected in wastewater samples are shaded in dark gray. The red dot represents the location of the WWTP where there was a positive detection of H5 (n=69), and the light blue dots represent participating WWTP where all samples were non-detect (n=78) during the study period. Alaska and Hawaii are not to scale.

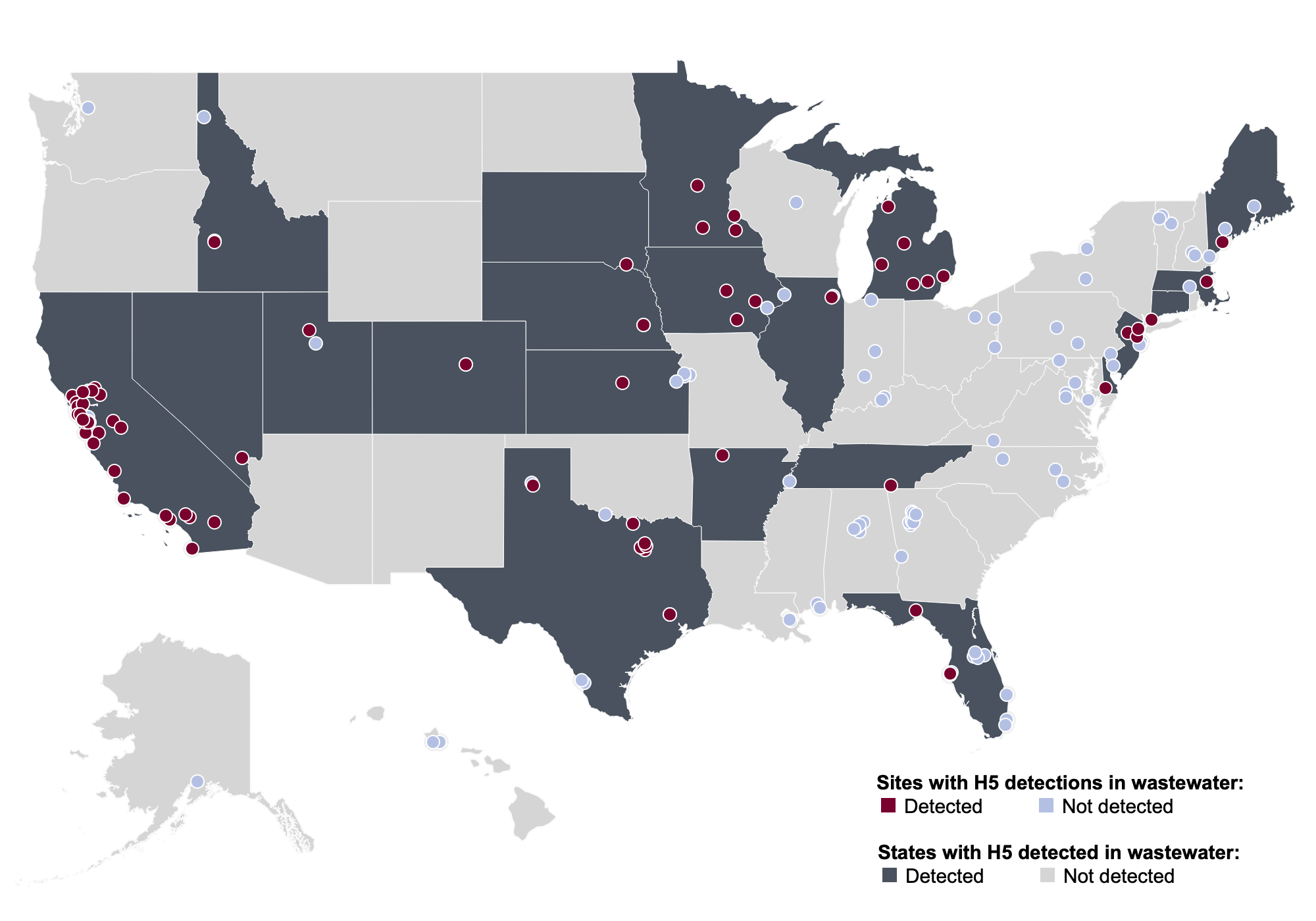

##### **Figure S2.** Wastewater sample positivity by region over time.

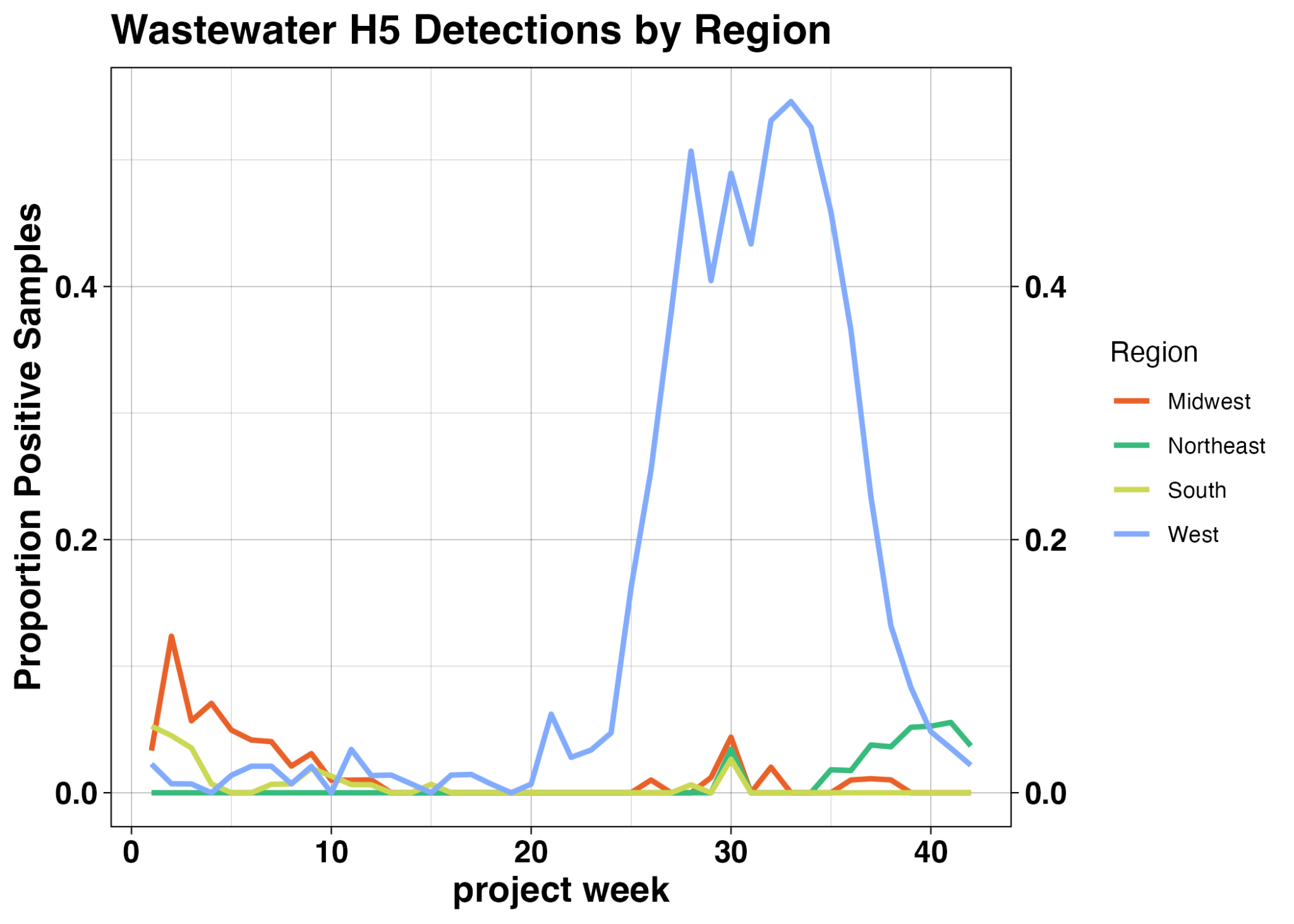

##### **Figure S3.** Wastewater samples positivity by state.

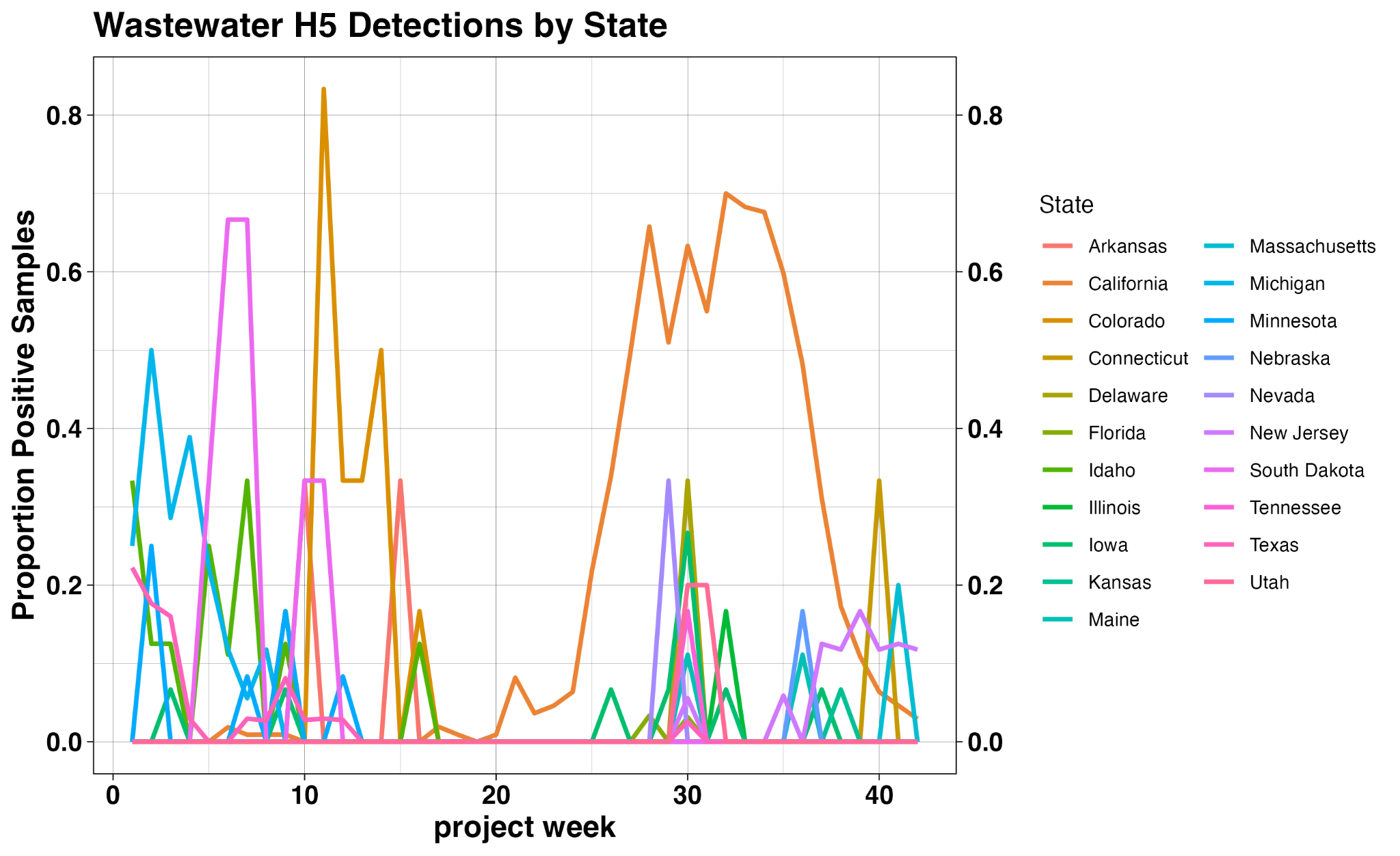

##### **Figure S4.** Total counts of positive herds, humans, poultry, wastewater samples, and wild birds recorded over the duration of the study, by region

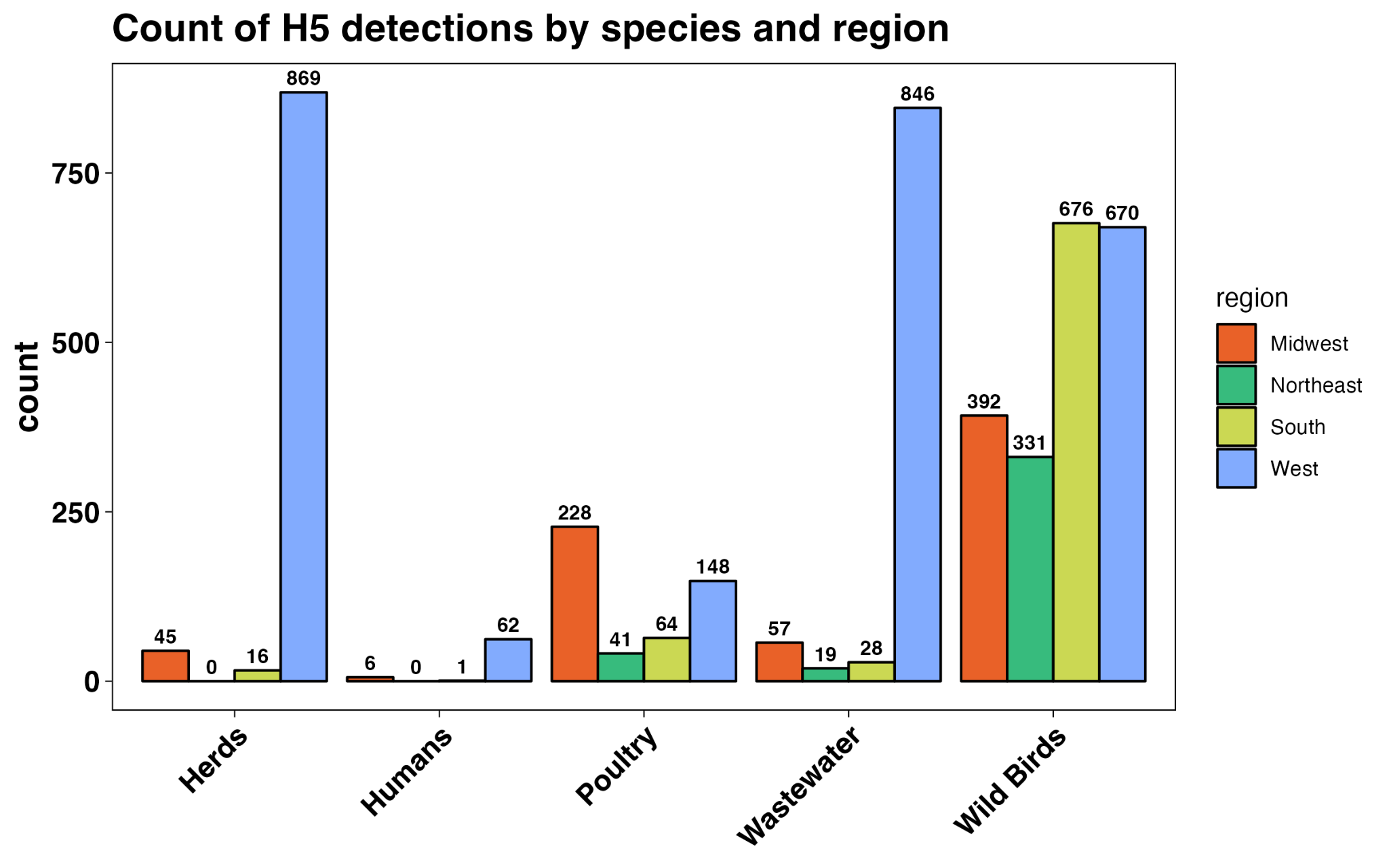

##### **Figure S5.** Correlation matrix for states with more than 10 detections of H5 in a WWTP during the study period (CA, CO, MI, NJ, TX).

P-value <0.05 = *, <0.01 = **, < 0.001 = ***.

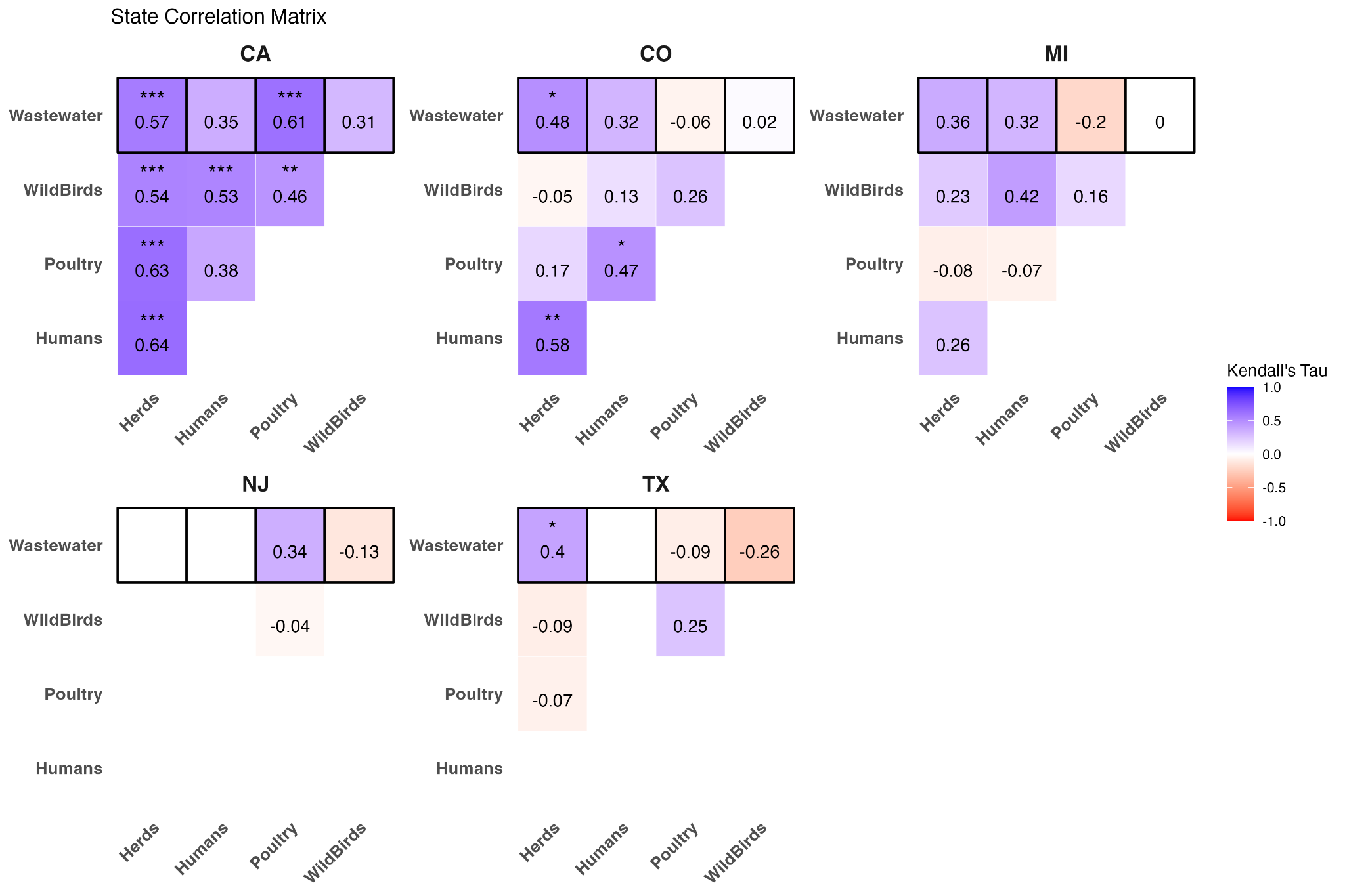
